# Five-year Transitions of Symptom Subtypes in Untreated Obstructive Sleep Apnea

**DOI:** 10.1101/2023.05.18.23290191

**Authors:** Jonna L. Morris, Paul W. Scott, Ulysses Magalang, Brendan T Keenan, Sanjay R. Patel, Allan I. Pack, Diego R. Mazzotti

**Author notes:** Corresponding Author: Jonna L. Morris, University of Pittsburgh, School of Nursing, 3500 Victoria Street, Room 420, Pittsburgh PA, 15261.

## Abstract

**Objectives:** It is unknown if symptom subtypes of obstructive sleep apnea (OSA) transition over time and what clinical factors may predict transitions.

**Methods:** Data from 2,643 participants of the Sleep Heart Health Study with complete baseline and 5-year follow-up visits were analyzed. Latent Class Analysis on 14 symptoms at baseline and follow up determined symptom subtypes. Individuals without OSA (AHI<5) were incorporated as a known class at each time point. Multinomial logistic regression assessed the effect of age, sex, body mass index (BMI) and AHI on specific class transitions.

**Results:** The sample consisted of 1,408 women (53.8%) and mean (SD) age 62.4 (10.5) years. We identified four OSA symptom subtypes at both baseline and follow-up visits: *minimally symptomatic, disturbed sleep, moderately sleepy* and *excessively sleepy*. Nearly half (44.2%) of the sample transitioned to a different subtype from baseline to follow-up visits; transitions to *moderately sleepy* were the most common (77% of all transitions). A five-year older age was associated with a 6% increase in odds to transit from *excessively sleepy* to *moderately sleepy* [OR (95% CI) = 1.06 (1.02, 1.12)]. Women had 2.35 times higher odds (95% CI: 1.27, 3.27) to transition from *moderately sleepy* to *minimal symptoms*. A 5-unit increase in BMI was associated with 2.29 greater odds (95% CI: 1.19, 4.38) to transition from *minimal symptoms* to *excessively sleepy*.

**Interpretation:** While over half of the sample did not transition their subtype over 5 years, among those who did, the likelihood of transitioning between subtypes was significantly associated with a higher baseline age, higher baseline BMI and with women, but was not predicted by AHI.

**Clinical Trials:** Sleep Heart Health Study (SHHS) Data Coordinating Center, (SHHS)https://clinicaltrials.gov/ct2/show/NCT00005275, NCT00005275

**Statement of significance:** There is very little research assessing symptom progression and its contributions to clinical heterogeneity in OSA. In a large sample with untreated OSA, we grouped common OSA symptoms into subtypes and assessed if age, sex, or BMI predicted transitions between the subtypes over 5 years. Approximately half the sample transitioned to a different symptom subtype and improvements in symptom subtype presentation were common. Women and older individuals were more likely to transition to less severe subtypes, while increased BMI predicted transition to more severe subtype. Determining whether common symptoms like disturbed sleep or excessive daytime sleepiness occur early in the course of the disease or as a result of untreated OSA over an extended period can improve clinical decisions concerning diagnosis and treatment.

---

Obstructive Sleep Apnea is a chronic and serious disorder that disrupts sleep, affects quality of life, and may result in increased risk for heart disease, metabolic disease, and dementia [1]. It is estimated that close to one billion people globally may have OSA[2]. Symptom reports, sleep architecture, medical comorbidities, anatomical characteristics, arousal threshold, loop gain, and social determinants of health have all been used to characterize OSA [3]. Symptom reports are clinically heterogeneous[3–12] They vary from those who report debilitating symptoms of excessive daytime sleepiness and insomnia to those who report minimal or no symptoms at all. The most common symptom reports or presentations may vary by population studied, for instance, women report more disturbed sleep symptoms than men [12–14] and older adults report impaired cognition [15]. Assessing and ascertaining phenotypes of OSA may help researchers and clinicians to better understand OSA heterogeneity, leading to improved diagnoses, knowledge of long-term health outcomes, and potentially, personalized treatment options.

Symptom subtypes of OSA have been extensively described [4,7,8,10–12,16] and have been shown to vary depending on disease severity and target sample. In studies of moderate to severe OSA (apnea-hypopnea index [AHI] ≥15 events/hour), the symptom subtypes of ‘disturbed sleep’, ‘minimal symptoms’, and ‘excessive daytime sleepiness’ (EDS) have been the most commonly identified [4,7,17]. Mazzotti et al. found an additional subtype of ‘moderate sleepiness’ in the Sleep Heart Health Study (SHHS) cohort [11]. Additional analyses in the SHHS showed that patients with mild OSA present with a similar symptom burden and subtype distributions as more severe disease [12]. Similar to prior research, relative to men, women were more likely to have disturbed sleep [3,5,18], while men comprised a greater proportion of the moderately sleepy group [12].

OSA symptom presentation is an important clinical indicator to inform screening and diagnosis [19]. However, little is understood about the natural course of the disease and how symptom presentation evolves in those with undiagnosed or untreated OSA. It is unclear whether symptoms of untreated OSA become more severe during the course of the disease. While it is frequently believed that symptoms worsen as the disease progresses over time, there are few data to support this contention. In fact, Zinchuk et al suggested that some individuals with OSA may become inured to their symptoms [3]. However, in others, OSA symptoms may significantly impair quality of life, daytime function, and severely impact mental well-being. As a result, the primary aim of this study is to enhance current knowledge of the presentation and 5-year change in OSA symptoms, as these are frequently the primary driving force for individuals to pursue diagnosis and treatment. The current study aimed to address this gap in a secondary analysis of SHHS data, by examining the proportion of participants that transitioned to a different symptom subtype over 5 years and determining whether baseline clinical and sociodemographic factors were associated with symptom transitions.

## Methods

### Study sample

Data used in the present study was collected as part of the SHHS, a multi-center community-based observational cohort that investigated the cardiovascular outcomes of untreated obstructive sleep apnea [20]. Participants (>40 years) completed their baseline assessments from 1995-1998 and follow up assessments five years later, between 2001 and 2003. Both assessments included clinical evaluations, self-reported questionnaires and at home polysomnography. Institutional Review Board approval was obtained at each investigational study site and each participant provided informed consent. Data are available from the National Sleep Research Resource (sleepdata.org)[21] Approval to conduct this study using data was granted from the National Sleep Research Resource. As all data were deidentified, institutional IRB approval was not necessary.

Sleep data was collected using polysomnography, as described in other publications [21]. An apnea was defined by a complete or near-complete cessation in breathing for at least 10 seconds and hypopneas were defined by a decrease in nasal airflow or chest or abdominal plethysmograph amplitude lasting for at least 10 seconds [20]. AHI was determined based on the number of apneas or hypopneas with ≥4% oxygen desaturation per hour of sleep [22]. OSA was defined based on an AHI≥5 events/hour.

### OSA symptom subtypes

Symptom subtypes at baseline and at the 5-year follow up visit were assessed in participants with OSA (AHI ≥5) using fourteen symptom items (see Appendix for list of symptoms), as previously described in the same cohort [11,12]. Questionnaires to assess symptoms included the Epworth Sleepiness Scale (ESS) [23], and specific questions from the Quality of Life Survey (SF 36) [24] and the Sleep Habits Questionnaire [25], which was developed for the parent study and captured symptoms such as snoring, waking up gasping for air, nighttime awakenings, and insomnia symptoms. OSA symptom subtypes were identified using latent class analysis (LCA) based on individual symptom items, as previously described [11,12].

### Statistical Analyses

Analyses were conducted using SAS 9.4 (SAS Institute, Cary, NC). Preliminary analyses included univariate and bivariate descriptive statistics on sample characteristics. LCA was performed separately at each visit (baseline and 5-year follow-up) amongst those with OSA. We opted for this method, instead of latent transition analyses to support consistency of symptom subtype membership observed in prior studies that looked at a single time point. We formally tested non-invariant and invariant models and Bayesian Information Criterion (BIC) results suggest strong preference for the invariant model (BIC=135023.1 vs. BIC=136424.9), supporting the use of LCA. The modal posterior probability of membership was used to assign individuals to symptom subtype clusters. The optimal solution for determining the number of subtypes was based on the BIC [26]. Symptom subtypes at baseline and follow-up were compared to assess the prevalence of transitions between groups over the study period. Multinomial logistic regression models were fit within each symptom subtype at baseline to examine the prediction of transitions to other subtypes at follow-up. Those without OSA (AHI<5 events/hour) at either baseline or follow-up were incorporated as having known group membership. Type 1 error inflation was controlled for by fitting predictors in a multinomial logistic regression model together. Those who stayed in the same group were treated as the reference in these models. We first fit models predicting transitions based on common risk factors for OSA, i.e., baseline age, sex, BMI, and AHI. We also examined how changes in BMI and AHI (except in analyses among those without OSA at baseline) between follow-up and baseline visits influenced transitions between groups. To facilitate interpretability, we report effect estimates for age as 5-year increments, BMI as 5-unit increments and AHI as 10-unit increments.

## Results

### Sample characteristics

Sample characteristics are listed in Table 1. Of the complete SHHS cohort, 5,804 men and women included in the baseline assessment and only 3,295 returned for the follow up assessment. In the current study, a total of 2,619 participants with available baseline and follow up AHI and symptom information were included. The total sample was balanced by sex (1,211 men [46.2%] and 1,408 women [53.8%]), with a mean ± standard deviation (SD) age of 62.4 ± 10.5 years. Participants were primarily White (87.2%), not Hispanic or Latino (95.5%), and overweight at baseline (BMI of 28.2 ± 4.9 kg/m^2^). They reported baseline ESS score of 7.8 ± 4.3 and baseline AHI of 9.3 ± 12.0 events/hour. Average time to follow up visit was 5.2 ± 0.3 years. The current study sample had a significantly lower mean AHI than the complete SHHS sample.

**Table 1.** Sample Characteristics at Baseline

| Variable | Category | Study Sample<br>(N=2,619) | Excluded<br>participants<br>(N=3,185) | P |
| --- | --- | --- | --- | --- |
|  |  | M (SD) for numeric;<br>N (%) for<br>categorical | M (SD) for<br>numeric;<br>N (%) for<br>categorical |  |
| Sex | Men | 1211 (46.2) | 1554 (48.8) | 0.05 |
|  | Women | 1408 (53.8) | 1631 (51.2) |  |
| Age, years | - | 62.4 (10.5) | 63.7 (11.7) | <0.01 |
| BMI, kg/m<sup>2</sup> | - | 28.2 (4.9) | 28.1 (5.2) | 0.39 |
| AHI, events/h | - | 9.3 (12.0) | 10.9 (14.8) | <0.01 |
| Race | Black | 178 (6.8) | 337 (10.6) | <0.01 |
|  | Other | 157 (6.0) | 225 (7.1) |  |
|  | White | 2284 (87.2) | 2623 (82.4) |  |
| Ethnicity | Hispanic or Latino | 119 (4.5) | 161 (5.1) | 0.37 |
|  | Not Hispanic or Latino | 2500 (95.5) | 3024 (95.0) |  |

AHI=Apnea-Hypopnea Index; BMI=body mass index; *Study sample* included participants with both baseline and follow up assessments; *Excluded participants* did not have a follow-up assessment

### Prevalence of OSA symptom subtypes is consistent at baseline and follow-up visits

Among participants with OSA at baseline, a four-class solution generated the best fit at each time point (baseline and follow-up) and yielded the most optimal solution based on the BIC. Latent classes were interpreted consistently across time points and in line with previous literature [11] as *disturbed sleep* (reports of low sleep latency, low sleep efficiency, and early waking), *excessively sleepy* (reports of remarkable sleepiness throughout the day and physical fatigue), *moderately sleepy* (falling asleep while watching TV or sitting quietly, but without the physical fatigue or reports of all day impairments), and *minimally symptomatic* (minimal reports of excessive daytime sleepiness or disturbed sleep). OSA symptom subtypes were similar in proportion between baseline and follow-up (Table 2), with the minimally symptomatic and moderately sleepy subtypes most prevalent. Predictors of transitions at baseline and follow-up by symptom subtype are shown in Table 3.

**Table 2.** Proportions of symptoms subtypes among those with OSA at baseline and follow-up visits.

| Time point | Disturbed Sleep | Excessively Sleepy | Moderately Sleepy | Minimally Symptomatic |
| --- | --- | --- | --- | --- |
| Baseline | 16.2% | 11.2% | 37.0% | 35.0% |
| Follow-up | 13.7% | 11.3% | 36.1% | 39.0% |

### Proportion of symptom subtype transitions over approximately five years

Figure 1 shows the proportion of each symptom subtype transition between baseline and follow-up. Approximately 56% of the cohort did not transition to a different subtype. Of those who transitioned, 77% transitioned to the moderately sleepy subtype. Of the transitions to the moderately sleepy subtype, 37.9% transitioned from the excessively sleepy subtype. Of those who were part of the moderately sleepy subtype at baseline, 20.6% transitioned to the minimally symptomatic subtype at follow up. Only 31.4% of those in the excessively sleepy subtype at baseline remained in that subtype after 5 years. From the minimal symptoms subtype, 19.8% transitioned to the moderately sleepy subtype at follow up. There were 19.4% who transitioned from the disturbed sleep subtype at baseline to the moderately sleepy subtype at follow up.

**Figure 1.**
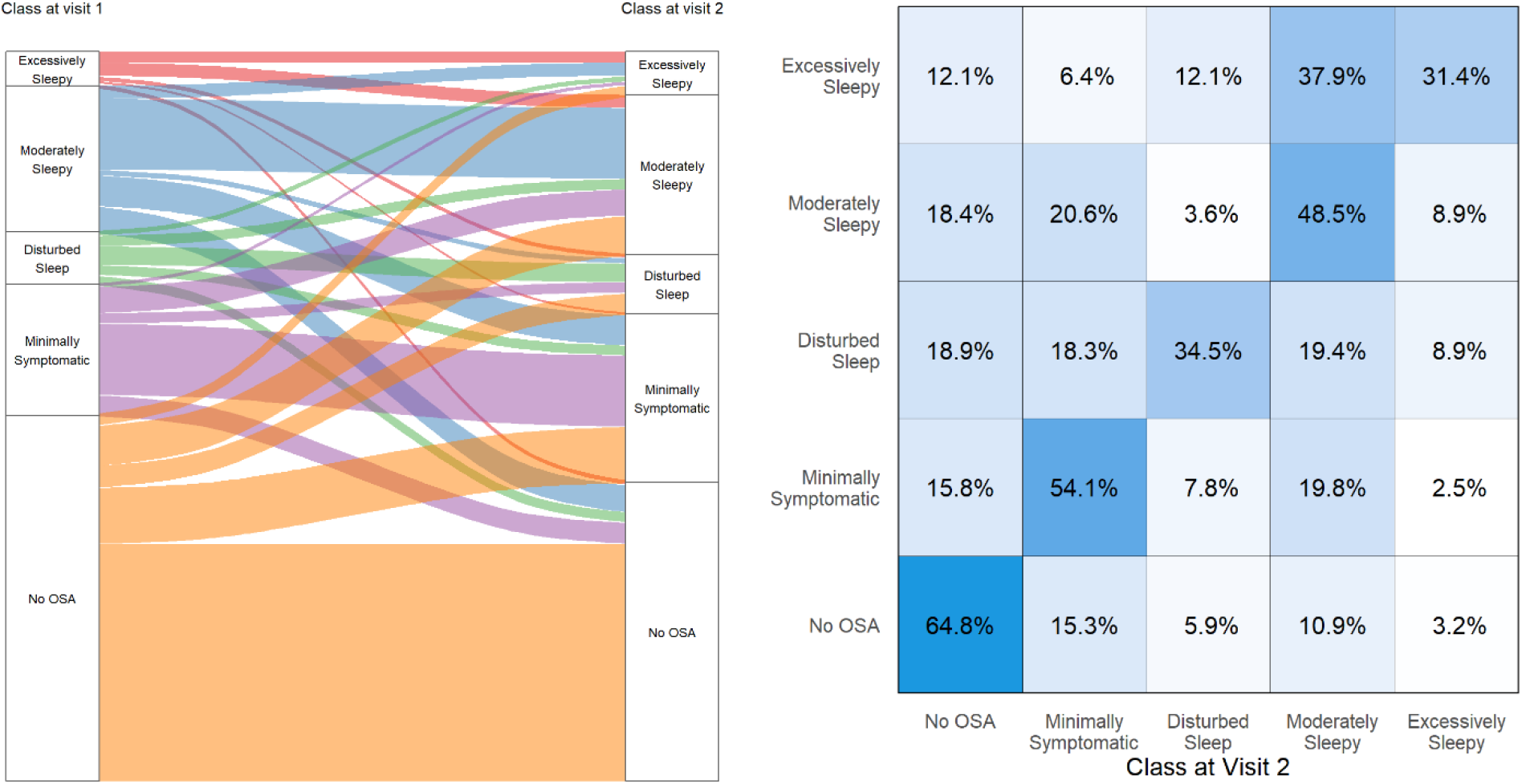
Proportions of each OSA symptom subtype transition between baseline and follow-up visits. The left plot shows the pattern of transitions relative to all participants. The right plot shows the proportions relative to each subtype at baseline. The diagonal boxes indicate the reference scenario where participants remained in the same symptom subtype. Brighter colors represent higher transition proportions.

### Predictors of symptom subtype transitions among those with OSA at baseline

Next, we determined whether baseline clinical and sociodemographic factors were associated with specific OSA symptom subtype transitions across the study period. Results of multinomial logistic regression assessing the association between baseline predictors and symptom subtype transitions among participants with OSA are summarized in Figures 2-6. Model specifications and results including all p-values are available in the supplementary tables.

**Figure 2a.**
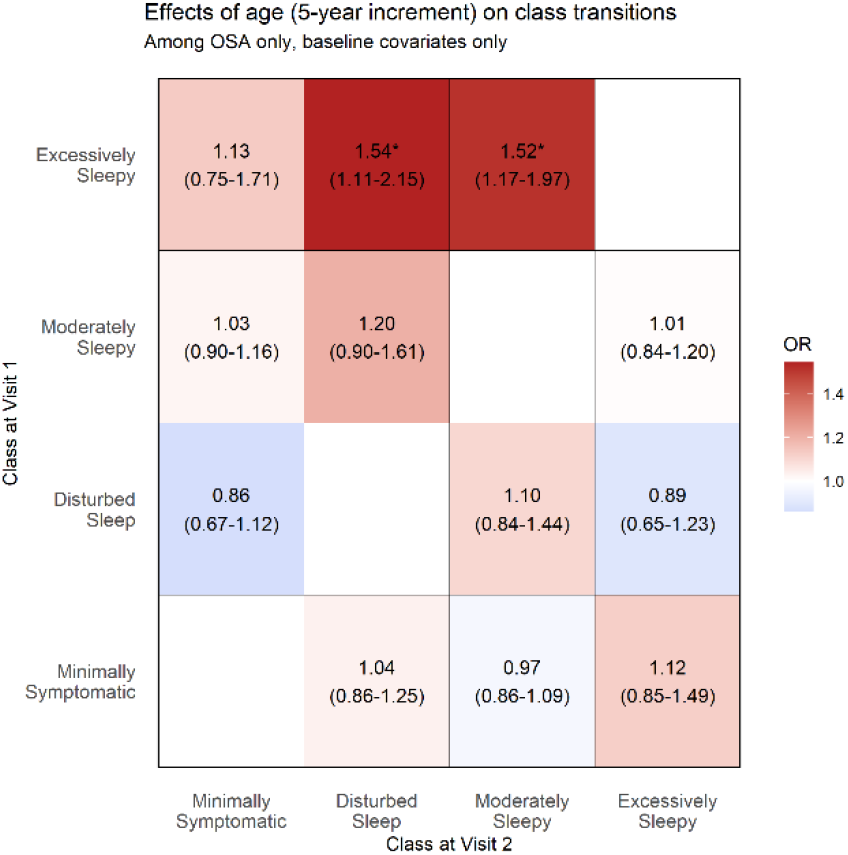
Figure 2a show the effects of age on class transitions from baseline, controlling for other baseline characteristics. The diagonal white boxes indicate the reference scenario where participants remained in the same symptom subtype. Brighter colors represent higher transition proportions. All colored boxes contain the odds ratio and confidence interval of the predictor of the transitions.

**Figure 2b.**
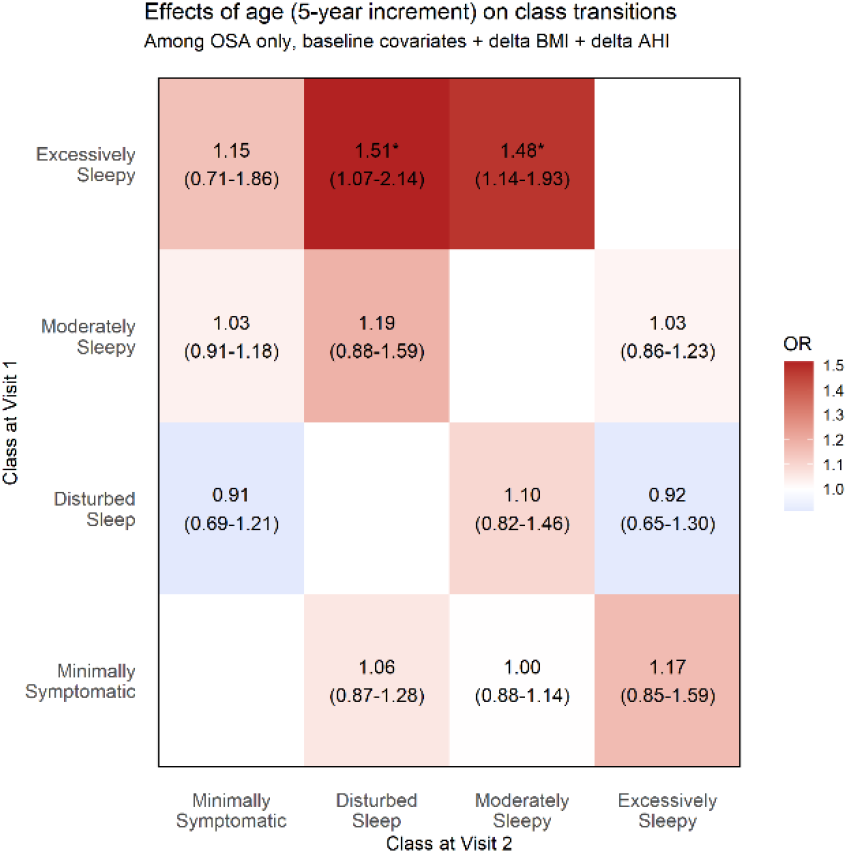
Figure 2b shows the effects of age on class transitions controlling for baseline plus changes in BMI and AHI. The diagonal white boxes indicate the reference scenario where participants remained in the same symptom subtype. Brighter colors represent higher transition proportions. All colored boxes contain the odds ratio and confidence interval of the predictor of the transitions.

**Figure 3a.**
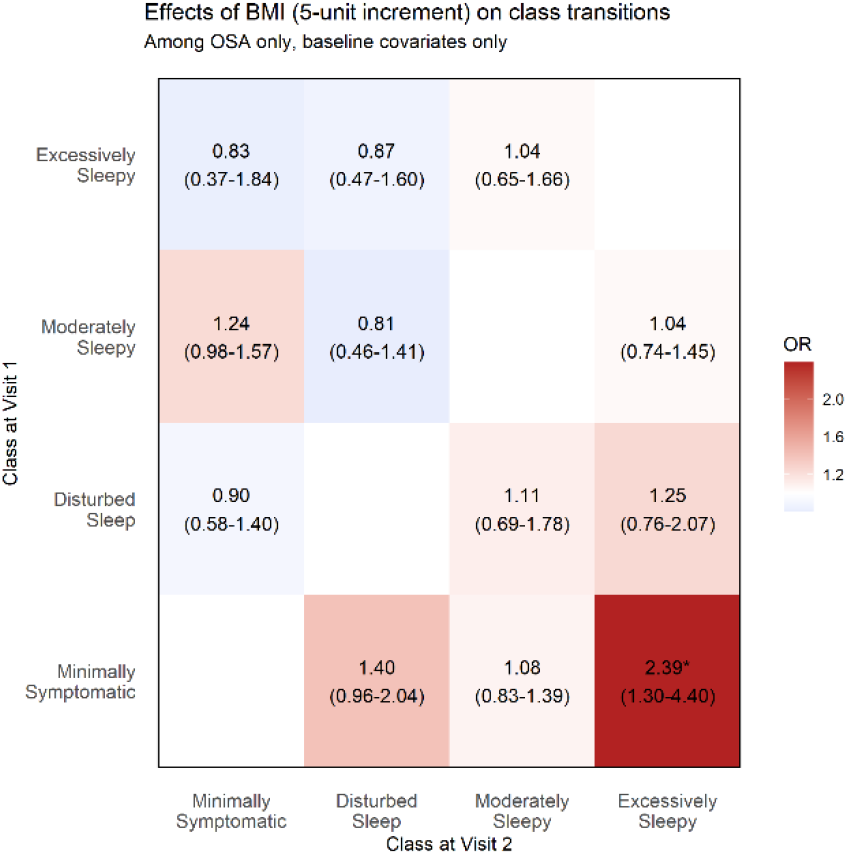
Figure 3a show the effects of BMI on class transitions from baseline, controlling for other baseline characteristics. The diagonal white boxes indicate the reference scenario where participants remained in the same symptom subtype. Brighter colors represent higher transition proportions. All colored boxes contain the odds ratio and confidence interval of the predictor of the transitions.

**Figure 3b.**
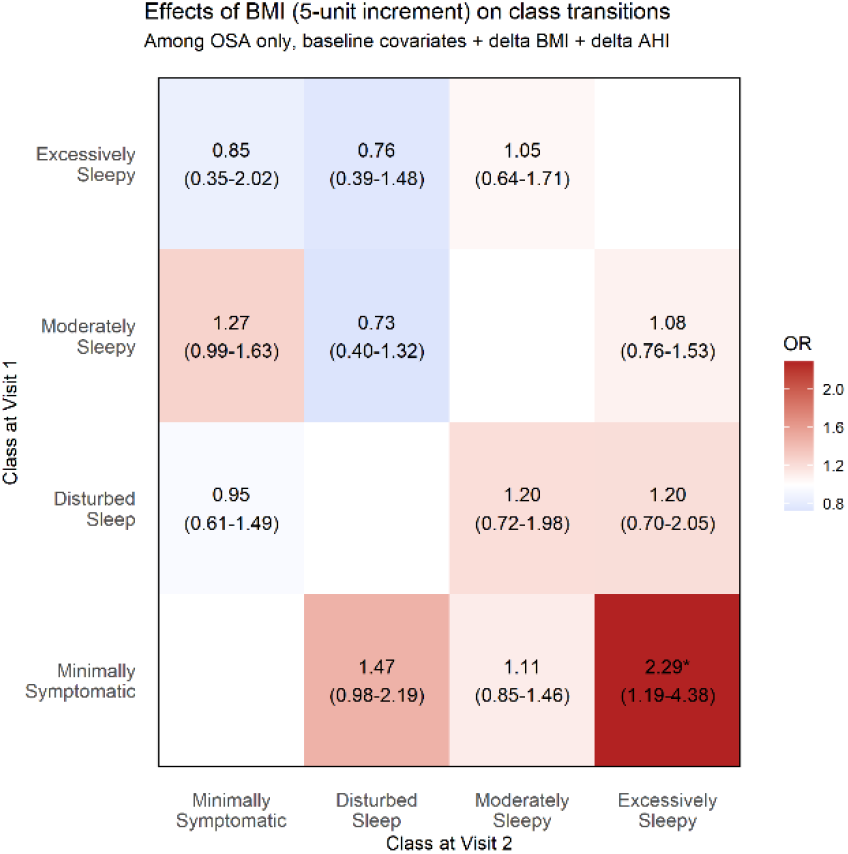
Figure 3b shows the effects of BMI on class transitions controlling for baseline plus changes in BMI and AHI. The diagonal white boxes indicate the reference scenario where participants remained in the same symptom subtype. Brighter colors represent higher transition proportions. All colored boxes contain the odds ratio and confidence interval of the predictor of the transitions.

**Figure 4a.**
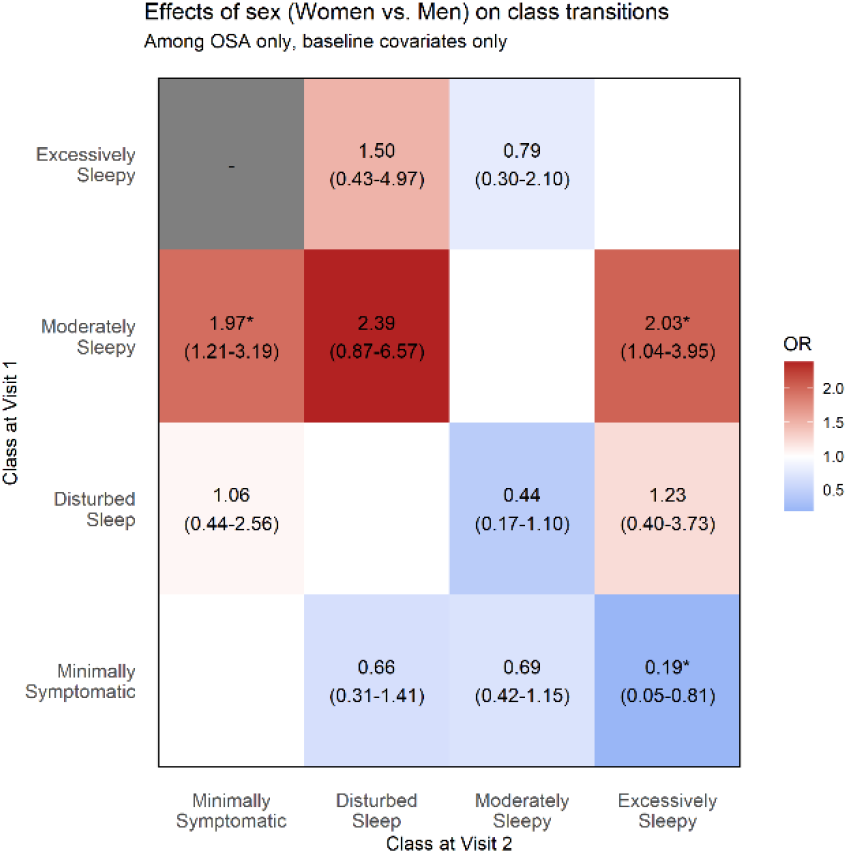
Figure 4a show the effects of sex on class transitions from baseline, controlling for other baseline characteristics. The diagonal white boxes indicate the reference scenario where participants remained in the same symptom subtype. Brighter colors represent higher transition proportions. All colored boxes contain the odds ratio and confidence interval of the predictor of the transitions.

**Figure 4b.**
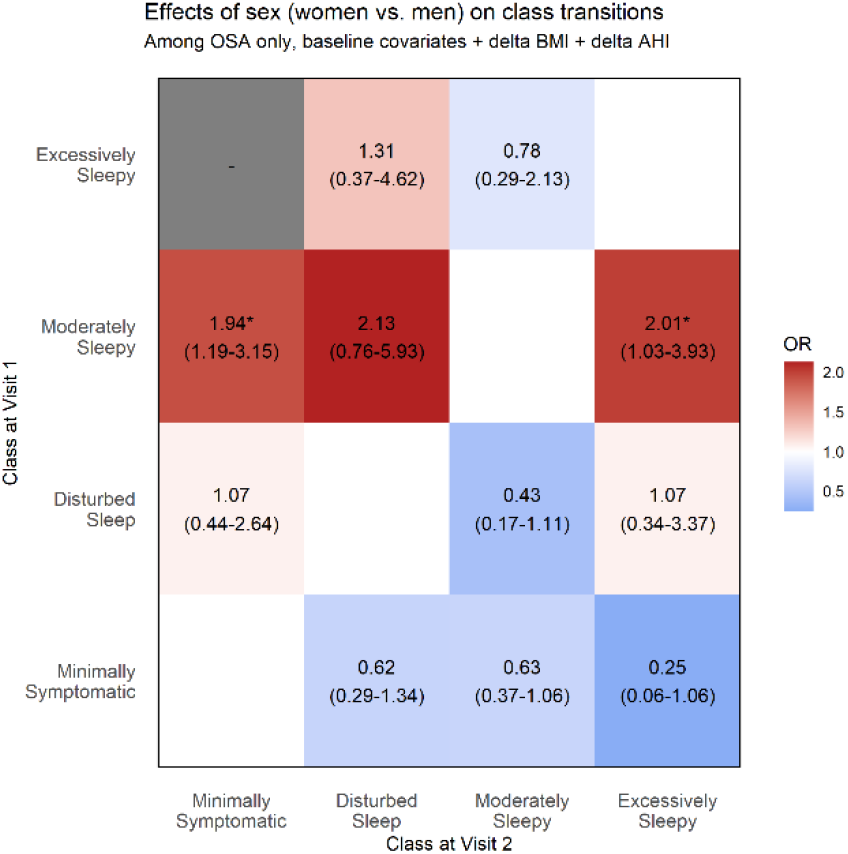
Figure 4b shows the effects of sex on class transitions controlling for baseline plus changes in BMI and AHI. The diagonal white boxes indicate the reference scenario where participants remained in the same symptom subtype. Brighter colors represent higher transition proportions. All colored boxes contain the odds ratio and confidence interval of the predictor of the transitions.

**Figure 5a.**
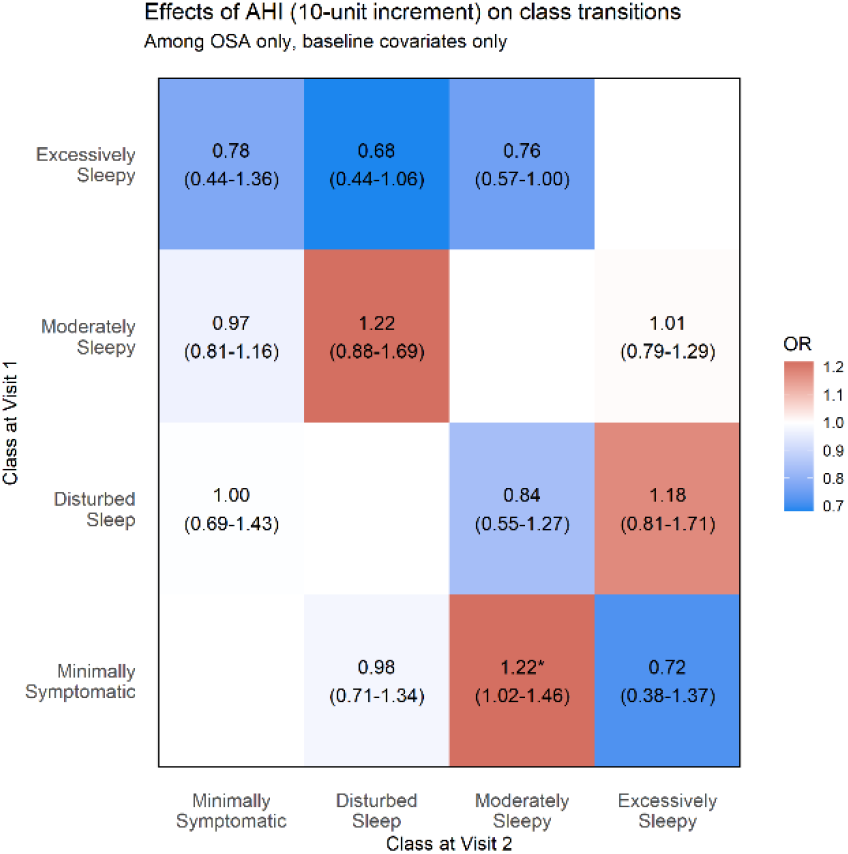
Figure 5a show the effects of AHI on class transitions from baseline, controlling for other baseline characteristics. The diagonal white boxes indicate the reference scenario where participants remained in the same symptom subtype. Brighter colors represent higher transition proportions. All colored boxes contain the odds ratio and confidence interval of the predictor of the transitions.

**Figure 5b.**
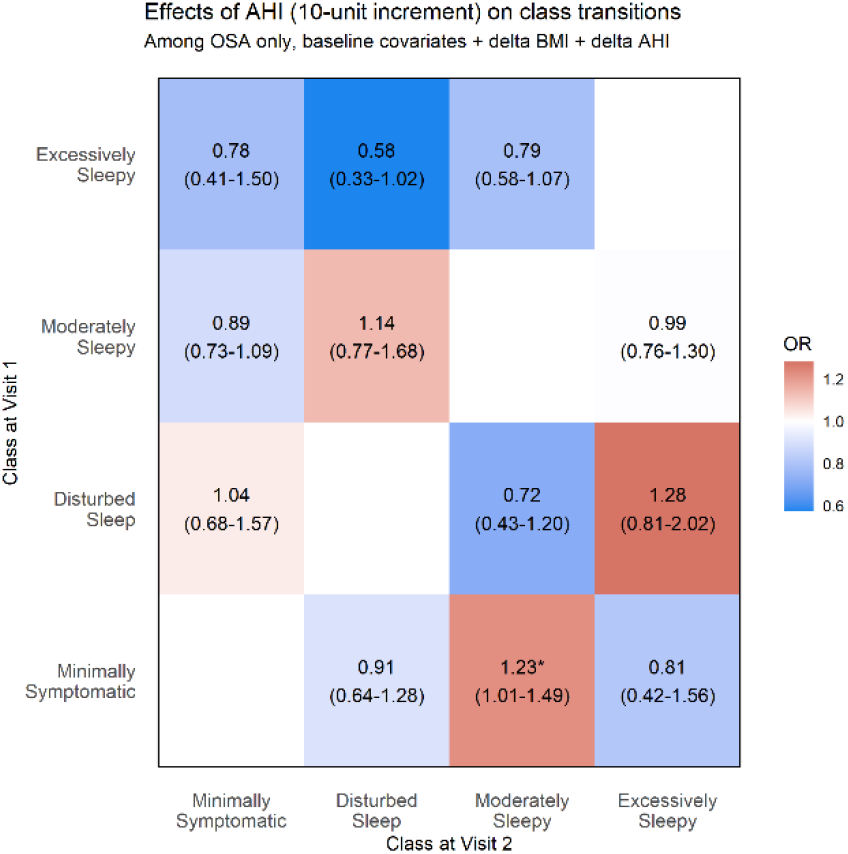
Figure5b shows the effects of AHI on class transitions controlling for baseline plus changes in BMI and AHI. The diagonal white boxes indicate the reference scenario where participants remained in the same symptom subtype. Brighter colors represent higher transition proportions. All colored boxes contain the odds ratio and confidence interval of the predictor of the transitions.

**Figure 6a.**
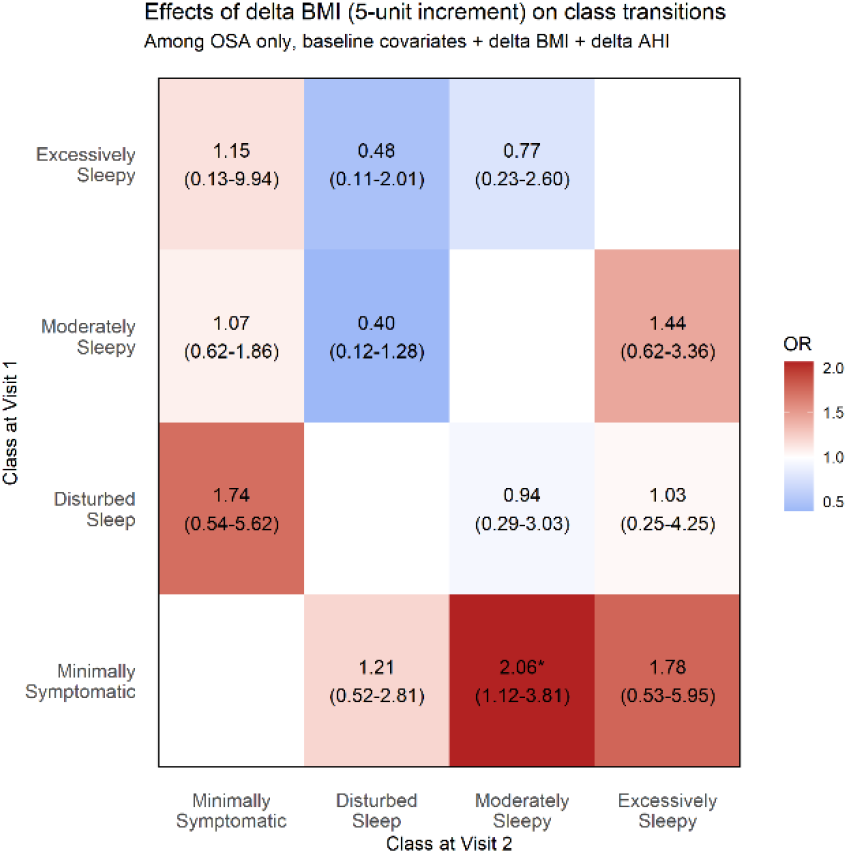
Figure 6a includes effects of the baseline covariates along with changes in BMI (delta BMI) and figure. The diagonal white boxes indicate the reference scenario where participants remained in the same symptom subtype. Brighter colors represent higher transition proportions. All colored boxes contain the odds ratio and confidence interval of the predictor of the transitions.

**Figure 6b.**
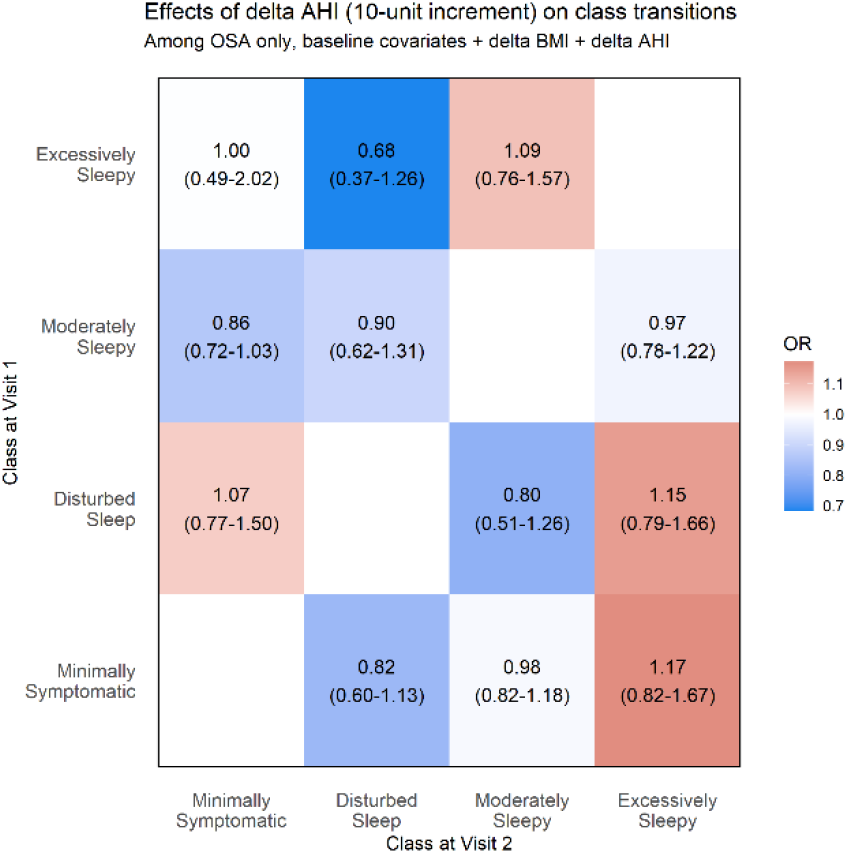
Figure 6b shows the effects of baseline covariates along with changes in AHI (delta AHI) among transitions from OSA symptom subtypes at baseline to the follow up visit. The diagonal white boxes indicate the reference scenario where participants remained in the same symptom subtype. Brighter colors represent higher transition proportions. All colored boxes contain the odds ratio and confidence interval of the predictor of the transitions.

Among participants who were excessively sleepy at baseline, there was a significant association between older age and transitioning to other subtypes, with a 5-year older age associated with a 1.54-fold increase in the odds of transitioning to the disturbed sleep subtype and as a 1.52-fold increase in the odds of transitioning to the moderately sleepy subtype. Both associations remained significant after adjusting for changes in BMI and AHI between baseline and follow-up. Age was not significantly associated with transitions from other subtypes at baseline.

Among participants who had minimally symptomatic OSA at baseline, a 5-unit increase in baseline BMI was associated with a 2.39-fold increase in the odds of transitioning to the excessively sleepy subtype (95% CI: 1.30, 4.40) controlled for baseline characteristics and a 2.29 fold-increase (95% CI: 1.19, 4.38) after additional adjustment for changes in BMI and AHI between the baseline and follow-up visit. BMI was not associated with transitions from other baseline subtypes.

Women with the minimally symptomatic subtype were five times less likely than men with the minimally symptomatic subtype to transition to the excessively sleepy subtype [OR (95% CI) = 0.19 (0.05, 0.81)]. In participants who belonged to the moderately sleepy subtype at baseline, women were at approximately 2-fold greater risk of transitioning to a different subtype, including a 2.03-fold increase in the odds of transitioning to the excessively sleepy subtype (95% CI: 1.04, 3.95) and a 1.97-fold increase in the odds of transitioning to the minimally symptomatic subtype (95% CI: 1.21, 3.19), as well as a non-significant 2.39-fold higher odds of transitioning to the disturbed sleep subtype (95% CI: 0.87, 6.57), compared to men at the follow-up. Associations remained similar after adjusting for changes in BMI and AHI between baseline and follow-up visit.

Among individuals of the minimally symptomatic subtype, for every 10 event/hour greater baseline AHI, there was 1.22-fold (95% CI: 1.02, 1.46) (baseline adjusted only) and 1.23-fold (95% CI: 1.01, 1.49) (adjusted for baseline predictors and change in BMI and AHI) increased odds of transitioning to the moderately sleepy subtype. AHI was not significantly associated with transitions from other subtypes.

Delta AHI was not significantly associated with any transitions. Among individuals of the minimally symptomatic subtype, there was a significant 2.06-fold increase in the odds of transitioning to the moderately sleepy subtype with a 5-unit increase in delta BMI.

### Predictors of OSA symptom subtype among those with incident OSA at 5-year follow-up

Supplemental Table 3 summarizes the associations between clinical and sociodemographic predictors and transitions from No OSA to OSA (e.g., incident cases of OSA at follow-up) of different symptom subtypes, relative to those that remained without OSA. A 5-year older age was significantly associated with a 1.30-fold increase in the odds of transitioning to OSA with a disturbed sleep subtype (95% CI: 1.2, 1.5), a 1.16-fold increase in the odds of transitioning to OSA with a minimally symptomatic subtype (95% CI: 1.1, 1.3) and a 1.18-fold increase in the odds of transitioning to OSA with a moderately sleepy subtype (95% CI: 1.1, 1.3). A 5-unit increase in BMI was associated with a 1.86-fold increase in the odds of transitioning to OSA with a disturbed sleep subtype (95% CI: 1.5, 2.4), a 1.52-fold increase in the odds of transitioning to OSA with a minimally symptomatic subtype (95% CI: 1.3, 1.8) and a 1.69-fold increase in the odds of transitioning to OSA with a moderately sleepy subtype (95% CI: 1.4, 2.1). Finally, women were 50% less likely to transition to OSA with a moderately sleepy subtype [OR (95% CI) = 0.51 (0.4, 0.7)]. Notably, transitions from No OSA at baseline to the excessively sleepy subtype of OSA at follow-up were not significantly associated with age, BMI, or sex, with a non-significant association between increased BMI [OR (95% CI) = 1.4 (0.9, 2.0)].

## Discussion

In the present study, we examined 5-year transitions among symptom subtypes in untreated OSA and their clinical predictors. In both the baseline and follow up cohorts, we identified the presence of four distinct symptom subtypes excessively sleepy, moderately sleepy, disturbed sleep, and minimally symptomatic. Our results demonstrate 56% of the sample did not transition their subtype after 5 years, suggesting that in the majority of OSA subjects, symptom presentation did not meaningfully change over time. Among the 44% who transitioned, it was common to transition from the excessively sleepy subtype to another subtype, with the majority transitioning to the moderately sleepy subtype. For instance, older age at baseline was associated with a greater likelihood of transitioning from the excessively sleepy subtype to either the disturbed sleep or moderately sleepy subtypes. Conversely, women were most likely to transition out of the moderately sleepy subtype at baseline primarily transitioning to the minimally symptomatic or excessively sleepy subtype at follow up. Baseline BMI was most associated with transitions to the sleepy subtypes. Baseline AHI was only significantly associated with the transition from the minimally symptomatic subtype to the moderately sleepy subtype and delta AHI was not associated with any transitions. Age, BMI, and sex did not significantly predict a transition from the No OSA subtype to the excessively sleepy subtype within five years of development of OSA.

Tempaku et al recently assessed symptom transitions in a Brazilian cohort of primarily untreated OSA patients over an 8-year period [27]. They incorporated clinical covariates such as age, sex, and BMI, as well as sleep architecture. They observed similar symptom subtypes to the present study including, ‘sleepy’, ‘minimal symptoms’, and ‘disturbed sleep’, but not a moderately sleepy subtype. Likewise, they also found that 45% of their sample transitioned to a different symptom subtype, while the majority of the sample did not. Finally, they also reported that a large portion (27%) of their sample transitioned from the ‘sleepy’ subtype to the ‘minimal symptoms’ subtype. While the Tempaku et al study identified AHI changes as a significant predictor of symptom subtype transitions, our findings suggest that AHI had a minimal influence on the transitions. Instead, we observed that transitions were primarily explained by age, sex, and BMI. Thus, while our study supports the findings of Tempaku et al regarding the proportions transitioning between subtypes, it provides alternative explanations for prediction of symptom subtype transitions over time.

Similar to Temaku et al, our findings also demonstrated significant transitions from the excessively sleepy to moderately sleepy or minimal symptoms subtypes suggesting a potential improvement in symptoms over time. For instance, of those who were in the excessively sleepy subtype at baseline, 38.9% transitioned to the moderately sleepy subtype at follow-up. Pien et al reported similar findings in a study of symptom subtype transitions in those who were treated with continuous positive airway pressure (CPAP) over 2 years [28]. Not surprisingly, participants treated with CPAP improved their symptom presentation, but they also found that the untreated group demonstrated an improved symptom presentation[28]. Thus, Zinchuk et al suggested that untreated OSA patients may become inured to their symptoms [3]. Habituation theories may support this supposition because it suggests a mechanism for the diminishing physiological or psychological response to a stimuli [29], such as excessive daytime sleepiness. Studies of chronic sleep deprivation in shift work disorder suggest that many people do not recognize, or have become desensitized to, excessive daytime sleepiness [30]. More research would be needed to determine if patients with OSA have a similar pattern.

Our findings of decreases in excessive daytime sleepiness associated with aging may also be a factor driving these improvements in daytime sleepiness. A meta-analysis of aging in OSA also reported similar findings, but the mechanism was unclear [31]. It is plausible that older adults may also become inured or more accepting of their symptoms. Or older adults who are retired may simply have greater opportunities to sleep, thus reducing symptoms of excessive sleepiness during the day. Alternatively, because older adults may need less sleep [32], they may experience less daytime sleepiness.

Obesity was significantly associated with transitions to moderate and excessive daytime sleepiness which may suggest obesity plays a direct role in OSA symptom severity. Obesity is considered a strong risk factor for OSA, as the excess weight causes fat to accumulate in the tongue leading to increased OSA severity [33]. However, obesity is also associated with excessive daytime sleepiness, with studies suggesting that excess body fat can lead to disrupted sleep patterns and increased daytime fatigue [34,35]. Studies that assessed differences of mean BMI among symptom subtypes have had conflicting findings [4,17], however our study clearly demonstrates a temporal effect of BMI on worsening symptom presentation that was not available in cross-sectional studies. Notably, the excessively sleepy subtype has been associated with incident cardiovascular disease in multiple studies [8,11,36], although not all [18,37]. Thus, overweight or obese individuals may benefit from weight management to prevent worsening symptoms and the potential risk for cardiovascular diseases.

In addition to effects with obesity and age, we found that, relative to men, women were less likely to be in the moderately sleepy subtype at baseline, as well as more likely to transition from the moderate sleepy subtype to another subtype at follow-up. The moderately sleepy type is differentiated from the excessively sleepy subtype because while it includes an endorsement of sleepy behaviors when sitting down or relaxing, it does not include such global impairments as not feeling rested in the morning, feeling sleepy throughout the day, and physical fatigue. Reasons why women may be less likely to exhibit the moderately sleepy subtype and more likely to transition out of this subtype remain to be determined, but could include factors such as sex differences in symptom reporting or menopausal status, or gendered reasons (i.e. caregiving responsibilities) that women do not prioritize, recognize, or report their daytime sleepiness.[38–40]

It is unknown how long anyone had OSA before their baseline visit. However, if the original sleep study was accurate, it could be inferred that individuals with incident OSA at the follow-up assessment had been experiencing it for a duration of less than 5 years. That the excessive sleepy subtype was not significantly predicted by age, sex, or BMI within the first 5 years may suggest that this symptom subtype evolves differently in OSA. The role of excessive daytime sleepiness on cardiovascular risk among patients with OSA is an active area of investigation [8,11,18,41] underscoring the importance of how understanding OSA disease and symptom progression may factor into long term health outcomes.

More research is needed to determine if the disease course of untreated OSA is correspondent to changes in symptoms and AHI consistently worsening over time or if it has a more complex progression.

Moreover, the assessment of short-term stability in OSA symptom presentation remains understudied and might help explain these findings.

Other factors to consider that may be contributing to symptom transitions are chronic or acute stress, recently developed chronic conditions such as type 2 diabetes, or a change in lifestyle or work hours. Further research will be needed to discern if the transitions are due to causes unrelated to OSA.

A little over half of the sample (56%) did not transition symptom subtypes with patients presenting with minimal symptoms at baseline being the least likely to transition to another subtype. Little is understood about patients diagnosed with OSA who report minimal symptoms except that they are less likely to adhere to continuous positive airway pressure treatment (CPAP) than patients who experience excessive daytime sleepiness [42,43]. With approximately half of patients who start CPAP failing to maintain adherence over the long-term [44], an understanding of which symptom subtypes remain stable or transition over time may assist in treatment decisions, particularly in the context of understanding the potential cardiovascular benefit of CPAP in asymptomatic individuals with OSA [45,46].

Our study also has important limitations. Sample size was limited to assess less prevalent transitions (e.g., lighter boxes in Figure 1, right panel) and the influence of other clinical covariates such type 2 diabetes, smoking or sleep characteristics. Studies to assess these clinical factors are also important and deserve attention in future studies. There may be selection bias in that the people who may have transitioned to excessive daytime sleepiness may have died. The symptoms measured to determine the LCA may not include all relevant OSA symptoms. For instance, nocturia, a common symptom of OSA was not captured in this study. It is also possible that the instruments used to measure the symptoms did not fully or accurately capture the symptoms experienced by the participants or may have been transient symptoms only reflecting the short time they were measured. A strength of the study is that while there is some uncertainty about CPAP status in the SHHS, prior literature indicates that only 2% may have been using CPAP [47,48], which is unlikely to substantially change the presented results.

In summary, this study represents preliminary work in assessing the evolution of symptoms in OSA over time. Our findings indicate that regardless of OSA disease severity, the majority of the sample had symptom subtypes that remained stable over time. While most health conditions tend to present with symptoms that worsen over time if left untreated (e.g., type 2 diabetes, cancer), our study suggests that many individuals with moderate sleepiness or even excessive sleepiness may improve their symptom presentation over the disease course, a counter intuitive finding. Accounting for clinical and demographic factors such as age, sex and BMI may assist clinicians with OSA treatment decisions. However, much more research is needed in additional cohorts, using different health measures to corroborate these findings.

## Data Availability

All data produced in the present study are available upon reasonable request to the authors

## Research/Grant Support

The Sleep Heart Health Study was supported by the NHLBI through the following cooperative agreements: U01HL53940 (University of Washington), U01HL53941 (Boston University), U01HL63463 (Case Western Reserve University), U01HL53937 (Johns Hopkins University), U01HL53938 (University of Arizona), U01HL53916 (University of California, Davis), U01HL53 934 (University of Minnesota), U01HL63429 (Missouri Breaks Research), and U01HL539 31 (New York University). The National Sleep Research Resource was supported by the NHLBI (HL114473). P01 HL094307 (Individual Differences in Obstructive Sleep Apnea).

## References

1. Veasey SC, Rosen IM. Obstructive Sleep Apnea in Adults. N Engl J Med. 2019;380(15):1442–1449. doi:10.1056/NEJMCP1816152/SUPPL_FILE/NEJMCP1816152_DISCLOSURES.PDF

2. Benjafield A V., Ayas NT, Eastwood PR, et al. Estimation of the global prevalence and burden of obstructive sleep apnoea: a literature-based analysis. Lancet Respir Med. 2019;7(8):687–698. doi:10.1016/S2213-2600(19)30198-5

3. Zinchuk A, Yaggi HK. Phenotypic Subtypes of OSA: A Challenge and Opportunity for Precision Medicine. Chest. 2020;157(2):403–420. doi:10.1016/j.chest.2019.09.002

4. Kim J, Keenan BT, Lim DC, Lee SK, Pack AI, Shin C. Symptom-Based subgroups of koreans with obstructive sleep apnea. J Clin Sleep Med. 2018;14(3):437–443. doi:10.5664/jcsm.6994

5. Keenan BT, Kim J, Singh B, et al. Recognizable clinical subtypes of obstructive sleep apnea across international sleep centers: A cluster analysis. Sleep. 2018;41(3). doi:10.1093/sleep/zsx214

6. Magalang UJ, Keenan BT. Symptom Subtypes in OSA: Ready for the Clinic? Chest. 2021;160(6):2003–2004. doi:10.1016/j.chest.2021.09.022

7. Ye L, Pien GW, Ratcliffe SJ, et al. The different clinical faces of obstructive sleep apnoea: A cluster analysis. Eur Respir J. 2014;44(6):1600–1607. doi:10.1183/09031936.00032314

8. Allen AJH, Jen R, Mazzotti DR, et al. Symptom subtypes and risk of incident cardiovascular and cerebrovascular disease in a clinic-based obstructive sleep apnea cohort. J Clin Sleep Med. 2022;18(9):2093–2102. doi:10.5664/jcsm.9986

9. Labarca G, Reyes T, Jorquera J, Dreyse J, Drake L. CPAP in patients with obstructive sleep apnea and type 2 diabetes mellitus: Systematic review and meta-analysis. Clin Respir J. 2018;12(8):2361–2368. doi:10.1111/crj.12915

10. Gagnadoux F, Le Vaillant M, Paris A, et al. Relationship between OSA clinical phenotypes and CPAP treatment outcomes. Chest. 2016;149(1):288–290. doi:10.1016/j.chest.2015.09.032

11. Mazzotti DR, Keenan BT, Lim DC, Gottlieb DJ, Kim J, Pack AI. Symptom subtypes of obstructive sleep apnea predict incidence of cardiovascular outcomes. Am J Respir Crit Care Med. 2019;200(4):493–506. doi:10.1164/rccm.201808-1509OC

12. Morris JL, Mazzotti DR, Gottlieb DJ, Hall MH. Sex differences within symptom subtypes of mild obstructive sleep apnea. Sleep Med. 2021;84:253–258. doi:10.1016/j.sleep.2021.06.001

13. Shepertycky MR, Banno K, Kryger MH. Differences between men and women in the clinical presentation of patients diagnosed with obstructive sleep apnea syndrome. Sleep. 2005;28(3):309–314. Accessed June 27, 2015. http://www.ncbi.nlm.nih.gov/pubmed/16173651

14. Wimms A, Woehrle H, Ketheeswaran S, Ramanan D, Armitstead J. Obstructive Sleep Apnea in Women: Specific Issues and Interventions. Biomed Res Int. 2016;2016:1764837. doi:10.1155/2016/1764837

15. Dunietz GL, Chervin RD, Burke JF, Conceicao AS, Braley TJ. Obstructive sleep apnea treatment and dementia risk in older adults. Sleep. 2021;44(9). doi:10.1093/SLEEP/ZSAB076

16. Labarca G, Dreyse J, Salas C, Letelier F, Jorquera J. A Validation Study of Four Different Cluster Analyses of OSA and the Incidence of Cardiovascular Mortality in a Hispanic Population. Chest. 2021;160(6):2266–2274. doi:10.1016/J.CHEST.2021.06.047

17. Keenan BT, Kim J, Singh B, et al. Recognizable clinical subtypes of obstructive sleep apnea across international sleep centers: A cluster analysis. Sleep. 2018;41(3). doi:10.1093/sleep/zsx214

18. Trzepizur W, Blanchard M, Ganem T, et al. Sleep Apnea–Specific Hypoxic Burden, Symptom Subtypes, and Risk of Cardiovascular Events and All-Cause Mortality. Am J Respir Crit Care Med. 2022;205(1):108–117. doi:10.1164/RCCM.202105-1274OC/SUPPL_FILE/DISCLOSURES.PDF

19. Kapur VK, Auckley DH, Chowdhuri S, et al. Clinical practice guideline for diagnostic testing for adult obstructive sleep apnea: An American academy of sleep medicine clinical practice guideline. J Clin Sleep Med. 2017;13(3):479–504. doi:10.5664/jcsm.6506

20. Quan SF, Howard B V., Iber C, et al. The Sleep Heart Health Study: Design, rationale, and methods. Sleep. 1997;20(12):1077–1085. doi:10.1093/sleep/20.12.1077

21. Zhang GQ, Cui L, Mueller R, et al. The National Sleep Research Resource: Towards a sleep data commons. J Am Med Informatics Assoc. 2018;25(10):1351–1358. doi:10.1093/jamia/ocy064

22. Berry RB et. al. The AASM Manual for the Scoring of Sleep and Associated Events: Rules, Terminology and Technical Specifications, Version 2.2. Itle.; 2015.

23. Johns MW. A new method for measuring daytime sleepiness: the Epworth sleepiness scale. Sleep. 1991;14(6):540–545. Accessed October 30, 2014. http://www.ncbi.nlm.nih.gov/pubmed/1798888

24. Ware JE, Sherbourne CD. The MOS 36-item short-form health survey (Sf-36): I. conceptual framework and item selection. Med Care. 1992;30(6):473–483. doi:10.1097/00005650-199206000-00002

25. Quan SF, Howard B V., Iber C, et al. The Sleep Heart Health Study: Design, rationale, and methods. Sleep. 1997;20(12):1077–1085. doi:10.1093/sleep/20.12.1077

26. Schwartz AR, Patil SP, Laffan AM, Polotsky V, Schneider H, Smith PL. Obesity and obstructive sleep apnea: pathogenic mechanisms and therapeutic approaches. Proc Am Thorac Soc. 2008;5(2):185–192. doi:10.1513/pats.200708-137MG

27. Tempaku PF, Oliveira e Silva L, Guimarães TM, et al. The reproducibility of clinical OSA subtypes: a population-based longitudinal study. Sleep Breath. 2022;26(3):1253–1263. doi:10.1007/s11325-021-02470-5

28. Pien GW, Ye L, Keenan BT, et al. Changing Faces of Obstructive Sleep Apnea: Treatment Effects by Cluster Designation in the Icelandic Sleep Apnea Cohort. Sleep. 2018;41(3). doi:10.1093/SLEEP/ZSX201

29. Rankin CH, Abrams T, Barry RJ, et al. Habituation revisited: An updated and revised description of the behavioral characteristics of habituation. Neurobiol Learn Mem. 2009;92(2):135–138. doi:10.1016/J.NLM.2008.09.012

30. Åkerstedt T, Wright KP. Sleep Loss and Fatigue in Shift Work and Shift Work Disorder. Sleep Med Clin. 2009;4(2):257–271. doi:10.1016/j.jsmc.2009.03.001

31. Iannella G, Vicini C, Colizza A, et al. Aging effect on sleepiness and apneas severity in patients with obstructive sleep apnea syndrome: a meta-analysis study. Eur Arch Oto-Rhino-Laryngology. 2019;276(3):3549–3556. doi:10.1007/s00405-019-05616-0

32. Li J, Vitiello M V., Gooneratne NS. Sleep in Normal Aging. Sleep Med Clin. 2018;13(1):1. doi:10.1016/J.JSMC.2017.09.001

33. Wang SH, Keenan BT, Wiemken A, et al. Effect of Weight Loss on Upper Airway Anatomy and the Apnea-Hypopnea Index. The Importance of Tongue Fat. Am J Respir Crit Care Med. 2020;201(6):718–727. doi:10.1164/RCCM.201903-0692OC

34. Vgontzas AN, Papanicolaou DA, Bixler EO, Kales A, Tyson K, Chrousos GP. Elevation of plasma cytokines in disorders of excessive daytime sleepiness: role of sleep disturbance and obesity. J Clin Endocrinol Metab. 1997;82(5):1313–1316. doi:10.1210/JCEM.82.5.3950

35. Vgontzas AN, Bixler EO, Tan TL, Kantner D, Martin LF, Kales A. Obesity without sleep apnea is associated with daytime sleepiness. Arch Intern Med. 1998;158(12):1333–1337. doi:10.1001/ARCHINTE.158.12.1333

36. Labarca G, Dreyse J, Salas C, et al. Risk of mortality among patients with moderate to severe obstructive sleep apnea and diabetes mellitus: results from the SantOSA cohort. doi:10.1007/s11325-020-02283-y/Published

37. Mehra R, Azarbarzin A. Sleep Apnea–Specific Hypoxic Burden and Not the Sleepy Phenotype as a Novel Measure of Cardiovascular and Mortality Risk in a Clinical Cohort. Am J Respir Crit Care Med. 2022;205(1):12–13. doi:10.1164/RCCM.202110-2371ED/SUPPL_FILE/DISCLOSURES.PDF

38. Luyster, F.S, Baniak, L.M, Chasens, E.R., Feeley, C.A., Imes, C.C., & Morris J. Sleep among working adults. In: Social Determinants of Sleep. Oxford University Press; 2019:119–138.

39. Morris JL, Rohay J, Chasens ER. Sex Differences in the Psychometric Properties of the Pittsburgh Sleep Quality Index. J Women’s Heal. 2017;00(00):jwh.2017.6447. doi:10.1089/jwh.2017.6447

40. Morris JL, Chasens ER, Brush LD. Gender as a principle of the organization of clinical sleep research. Nurs Outlook. Published online August 1, 2020. doi:10.1016/j.outlook.2020.06.006

41. Eulenburg C, Celik Y, Redline S, et al. Cardiovascular Outcomes in Adults with Coronary Artery Disease and Obstructive Sleep Apnea with vs without Excessive Daytime Sleepiness in the RICCADSA Cohort. https://doi.org/101513/AnnalsATS202208-676OC. Published online February 17, 2023. doi:10.1513/ANNALSATS.202208-676OC

42. Weaver TE, Mancini C, Maislin G, et al. Continuous positive airway pressure treatment of sleepy patients with milder obstructive sleep apnea: results of the CPAP Apnea Trial North American Program (CATNAP) randomized clinical trial. Am J Respir Crit Care Med. 2012;186(7):677–683. doi:10.1164/rccm.201202-0200OC

43. Weaver TE, Chasens ER. Continuous positive airway pressure treatment for sleep apnea in older adults. Sleep Med Rev. 2007;11(2):99–111. doi:10.1016/j.smrv.2006.08.001

44. Bakker JP, Weaver TE, Parthasarathy S, Aloia MS. Adherence to CPAP: What Should We Be Aiming For, and How Can We Get There? Chest. 2019;155(6):1272–1287. doi:10.1016/j.chest.2019.01.012

45. Ryan S. Pro: should asymptomatic patients with moderate-to-severe OSA be treated? Breathe. 2019;15(1):7. doi:10.1183/20734735.0340-2018

46. Vakulin A, Chai-Coetzer CL, Mcevoy RD. Con: should asymptomatic patients with moderate-to-severe OSA be treated? Breathe. 2019;15:11–14. doi:10.1183/20734735.0347-2018

47. Gottlieb DJ, Yenokyan G, Newman AB, et al. Prospective study of obstructive sleep apnea and incident coronary heart disease and heart failure: the sleep heart health study. Circulation. 2010;122(4):352–360. doi:10.1161/CIRCULATIONAHA.109.901801

48. Redline S, Yenokyan G, Gottlieb DJ, et al. Obstructive sleep apnea-hypopnea and incident stroke: the sleep heart health study. Am J Respir Crit Care Med. 2010;182(2):269–277. doi:10.1164/RCCM.200911-1746OC

